# State-level RN density and age-adjusted covid-19 mortality: Contribution of the nursing workforce to pandemic response

**DOI:** 10.64898/2026.01.25.26343723

**Authors:** Rigan Louis, S. Nazmus Sakib, Pei Qinglin, Leslie A. Parker, J. Glenn Morris

## Abstract

Nurses represent the largest segment of the United States healthcare workforce and played an instrumental role in the country’s response to the COVID-19 pandemic. Yet, little attention has been given to the contribution of this component of the U.S. medical personnel in the nation’s ability to face public health crisis. We present a cross-sectional, ecological analysis using cumulative annual reports from different national databases to assess the relationship between registered nurse (RN) density at a state level and age-adjusted COVID-19 mortality within the state, using data from 2021 when mortality rates were peaking in the U.S.

At the state level, an increase of 1,000 RNs per 100,000 people, was associated with an estimated 24 to 44 fewer COVID-19 deaths per 100,000 residents (B= -0.024, *β*= -0.146, 95% CI: -0.044 to -0.003, p = .024). In this multivariate analysis including medical co-morbidities, vaccination, health insurance, and poverty level, RN density explained nearly 11% of the variability in COVID-19 mortality among states. Our findings underscore the critical role played by nurses in responding to the COVID-19 pandemic, and the importance of incorporating nursing workforce data into planning for future public health emergencies.

## 1. Introduction

Coronavirus disease-2019 (COVID-19), caused by the severe acute respiratory syndrome coronavirus 2 (SARS-CoV-2), has been one of the most devastating pandemics since the Spanish flu(1). As of September 2025, over 778.5 million people have been infected, and an estimated 7.1 million people have died from COVID-19 worldwide, including nearly 1.2 million deaths in the US alone, representing nearly 17% of the global death toll(2–4). The highest annual cumulative COVID-19 mortality in the US was observed in 2021(5). This observation is arguably due to several factors, including the introduction of Delta (and, to a lesser extent, Omicron) variants, vaccination mistrust and hesitancy, and longstanding socioeconomic disparities(6),(7,8). Moreover, COVID-19 has exacerbated preexisting issues affecting the US healthcare workforce and exposed systemic shortcomings in the country’s ability to respond to a public health crisis.

The medical and public health response to COVID involved multiple occupational groups within the healthcare workforce, including nurses. Nurses play a pivotal role in healthcare delivery by providing approximately 80% of hands-on primary care and acting as frontline healthcare personals(9). Yet, little attention has been given to their critical role in responding to public health crises, including the COVID-19 pandemic(10). There are an estimated 29 million nurses globally, representing nearly half of the world healthcare workforce(11,12). In the US, nursing is the largest healthcare profession, with an estimated 5 million licensed professionals, which include registered nurses (RNs), practical/vocational nurses (LPNs/LVNs), and advanced practice registered nurses (APRNs)(13). Despite this large body of professionals, the US nursing workforce remains understaffed due to multi-layered internal systemic challenges and additional extrinsic factors(14). With an average age of 48 years old, a significant segment of the nursing workforce is approaching retirement(15). Nurses’ distribution varies significantly by US states, with some states experiencing severe shortages while others are oversupplied(16,17). State-level registered nurse density, defined as the number of active RNs for a given state, divided by that state’s population, also differs widely across U.S. states(18). These disparities in state-level RNs density may have various negative implications for both nurses and the U.S. population, including reduced healthcare access, nurse burnout, and suboptimal patient care(19–21).

Several lines of evidence support a relationship between inadequate nurse-to-patient ratio and increased risk of adverse patient outcomes, including patients’ death(21–24). However, the relationship between state-level RNs density and adverse health outcomes in the context of COVID-19 mortality has not been well studied. This study aims to assess the association between state-level density and age-adjusted COVID-19 mortality in the U.S. in 2021, data which serve as a key component of efforts to optimize preparation for possible future pandemics.

## 2. Materials and Methods

### 2.1. Study design and Data Sources

This ecological, cross-sectional exploration uses publicly available cumulative annual reports from different national databases for 2021. All the 50 U.S. states and the District of Columbia (DC) were included in this study. State-level age-adjusted COVID-19 mortality (number of deaths per 100,000 people), was defined as the cumulative weighted averages of the age-specific death rates attributed to COVID-19 and was imported from CDC WONDER(25). State-level RN density, or number of RNs per 100,000 people, was calculated using data reported by the U.S. Bureau of Labor Statistics and US Census Bureau for 2021(16,26). State-level percentage of comorbidities (or chronic health conditions), defined as the average percentage of the population with at least one chronic health condition for each individual state, was estimated using data from the CDC’s Behavioral Risk Factor Surveillance System survey(27). To estimate state-level percentage of comorbidities, we pooled together state-level estimates from five different chronic health conditions, namely chronic obstructive pulmonary disease among adults, diabetes among adults, adults with obesity, adults with asthma, and 65-year-old and older adults hospitalized with heart failure as a principal diagnosis. In addition, state-level percentage of the population who have completed at least one series of COVID-19 vaccine and percentage of the population with health insurance coverage were obtained, respectively, from the U.S. Department of Health & Human Services and the U.S. Census Bureau(28,29). Percentage of the population living below 150% of the poverty threshold was used as a proxy of poverty index across the US states; the data was obtained from the CDC and U.S. Census’s American Community Survey(ACS) (30,31).

### 2.2. Variables

State-level age-adjusted COVID-19 mortality was used as the dependent variable, while RN density was the independent variable. The four other variables were used as covariates in the adjusted model. These covariates were carefully chosen based on their clinical relevance and from prior studies. They were limited to four to ensure the model’s stability, prevent any risk of overfitting, and optimize reliable findings.

### 2.3. Statistical Analysis

No sample size calculation was formally conducted given that this study was limited to the U.S. states and DC. All the data were analyzed as continuous variables. Means with their confidence intervals and standard deviations, as well as minimum and maximum values for each variable, were estimated for descriptive purposes. In addition, two maps were created to visualize the spatial distribution of respectively RN density and COVID-19 mortality across the US. The CDC’s BRFSS survey data for 2021 reported no data for the state of Florida for four of the five chronic health conditions, namely diabetes, COPD, obesity, and asthma. Estimates from 2020 and 2022 were averaged and used to replace these missing values in this analysis. Following the assessment of appropriate assumptions such as linearity, homoscedasticity, and normality of the residuals, initial univariate linear models were fitted to assess the relationship between COVID-19 mortality and each individual independent variable. Subsequently, a multivariate model was fitted using Ordinary Least Square (OLS) regression method, followed by sensitivity analysis. Stepwise analysis was performed to confirm whether all these covariates should be kept in the final model. A *p-value* ≤ .05 was indicative of statistical significance. ArcGIS Pro (version 3.3.0, 2024) hosted under the institutional license provided by the University of Florida was utilized to create maps. The basemap for the US was exported from the GADM repository(32). RStudio(33) version 2025.05.1 was used to perform all necessary analyses.

## 3. RESULTS

### 3.1. Descriptive Statistics

COVID-19 mortality was less pronounced among the Northern states of the US during the study period (Fig 1), while states located in the southwestern region of the US had the lowest RN density (Fig 2). Age-adjusted COVID-19 mortality varied across the US in 2021, with a national average of 99 (95%CIs: 89 to 108) deaths per 100,000 people.

**Fig 1.**
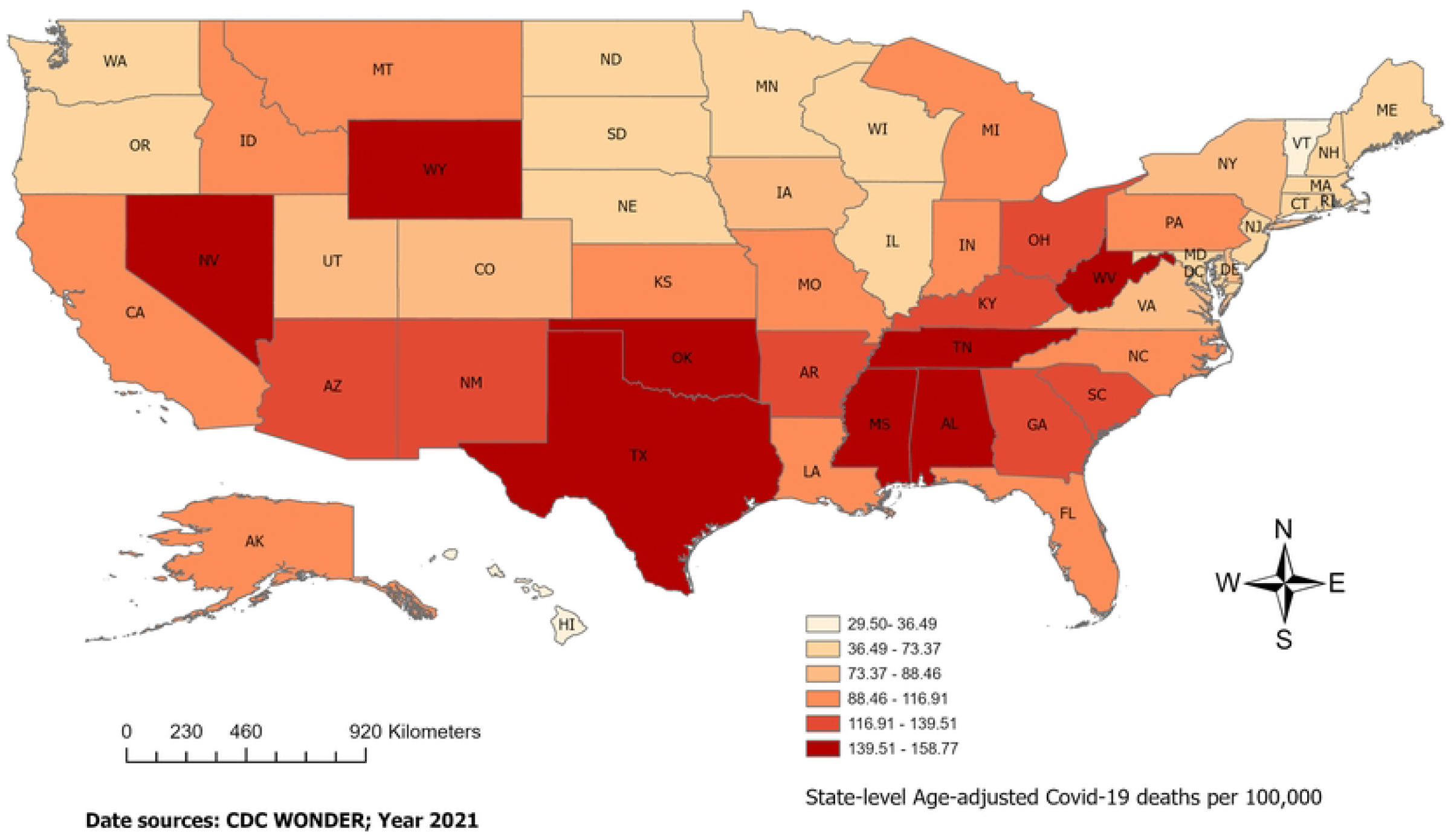
Age-Adjusted COVID-19 mortality distribution across US states in 2021. The Northern states showed a lower rate of cumulative COVID-19 mortality in 2021.

**Fig 2.**
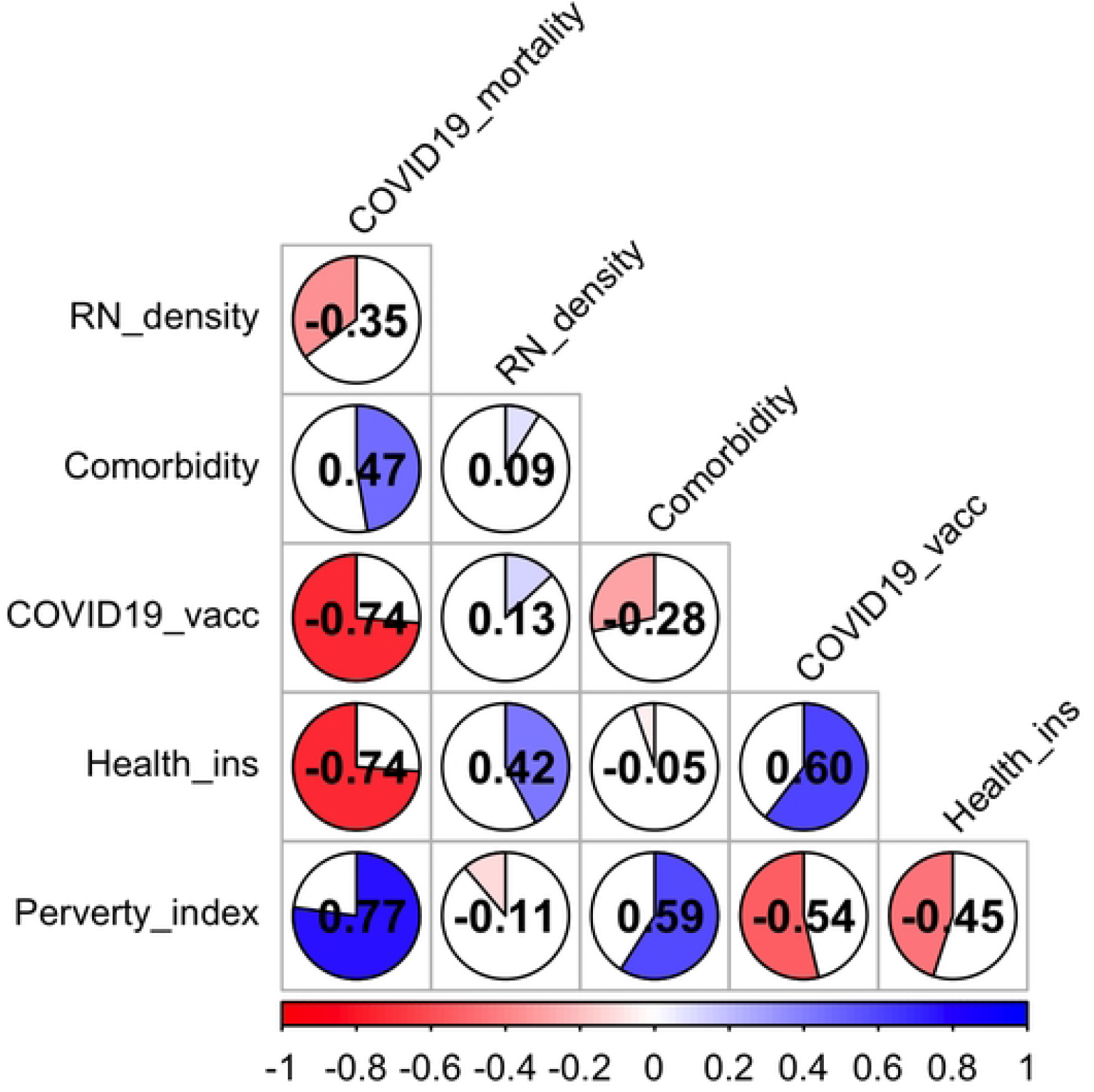
RN density distribution across the US in 2021. States in the southwestern region of the US had the lowest RN density 1n 2021.

The states of Vermont and Oklahoma had reported, respectively, the lowest and the highest death rates, thirty deaths per 100,000 people and 159 deaths per 100,000 people (Table 1). The estimated national average of RN density during the study period in the US was 975 RNs per 100,000 (or 9 RNs per 1,000) people. At the state level, RN density ranged from 712 RNs per 100,000 people in Utah to 1,579 per 100,000 people in South Dakota. RN density was higher than all the U.S. states in (1,722 per 100,000 people).

**Table 1.** Descriptive statistics.

| Variable | Mean | SD | CI Lower | CI Upper | Min | Max |
| --- | --- | --- | --- | --- | --- | --- |
| COVID-19 mortality | 99 | 34 | 89 | 108 | 30 | 159 |
| RN density | 975 | 209 | 917 | 1034 | 712 | 1722 |
| Comorbidity | 17 | 2 | 16 | 17 | 13 | 22 |
| COVID-19 Vacc. | 60 | 8 | 58 | 63 | 46 | 78 |
| Health insurance | 92 | 3 | 91 | 93 | 82 | 98 |
| Poverty index | 20 | 4 | 19 | 21 | 12 | 31 |

CI = Confidence interval; All estimates were calculated in R-studio; COV19 = Covid-19; Poverty index = Percentage of the population living below 150% of the poverty threshold.

State-level percentage of health insurance coverage and percentage of population who have completed at least a series of COVID-19 vaccine were strongly and negatively correlated with COVID-19 mortality, followed by RN density, which displayed a moderate correlation (Fig 3). State-level percentage of comorbidities and percentage of population living below 150% of the poverty threshold (Poverty index) showed, respectively, a moderate and a strong positive correlation with COVID-19 mortality (Fig 2).

**Fig 3.**
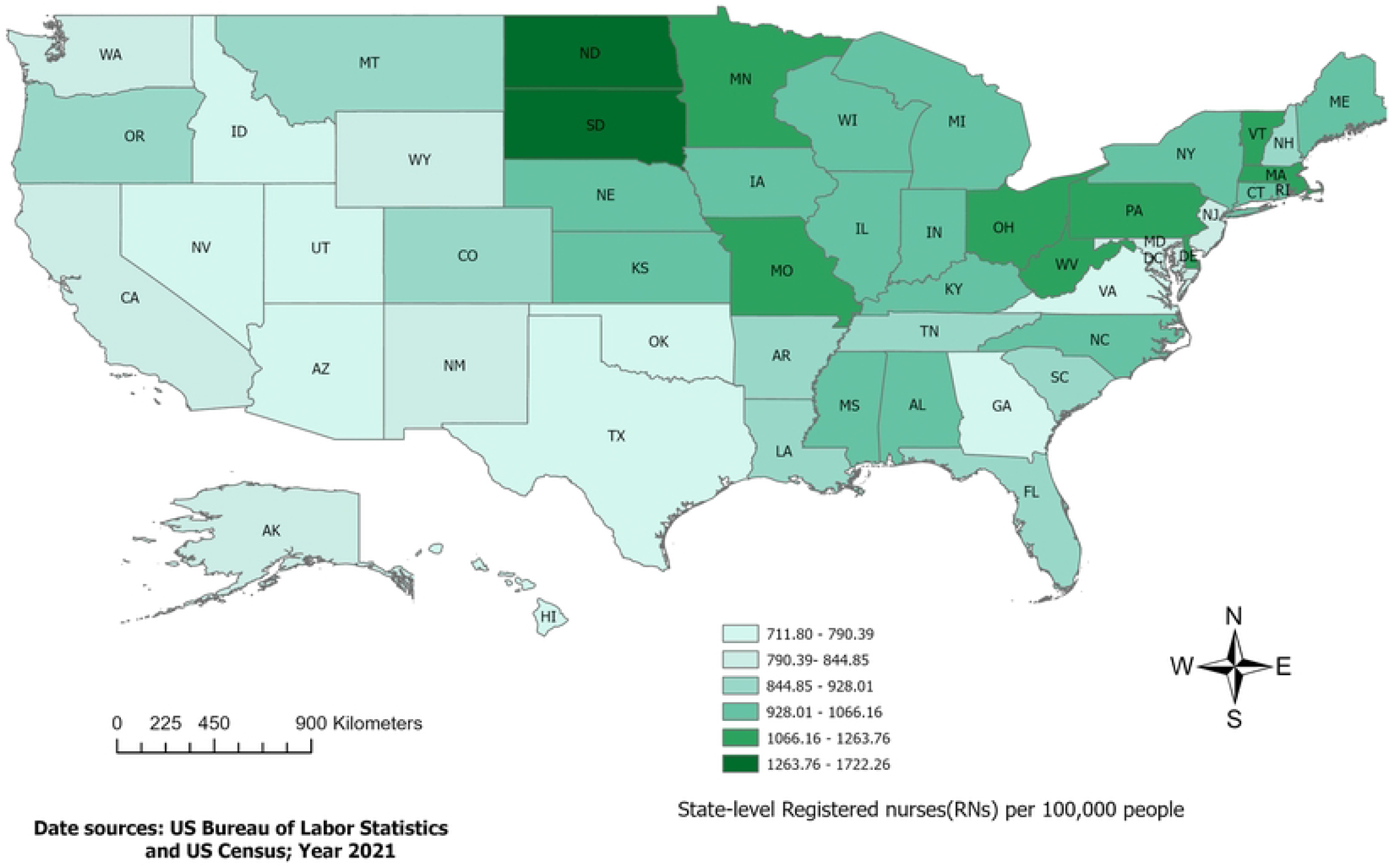
Correlation among the variables. Blue pie charts indicate positive correlation, and the red-to-pink show negative correlations.

### 3.2. Simple Linear Regression Results

Simple linear regression showed a negative statistically significant association between RN density and age-adjusted COVID-19 mortality, at the state level. An increase of 1,000 RNs per 100,000 people, was associated with an estimated 57-100 fewer COVID-19 deaths per 100,000100K residents (B= -0.057, 95% CI: -0.100 to -0.013, p = .012). Meanwhile, RN density only explained 12% of the variance of COVID-19 mortality across the US states and DC. The four covariates, individually, explained a moderate to high portion of the variance of COVID-19 mortality in the crude analyses (Fig 4).

**Fig 4.**
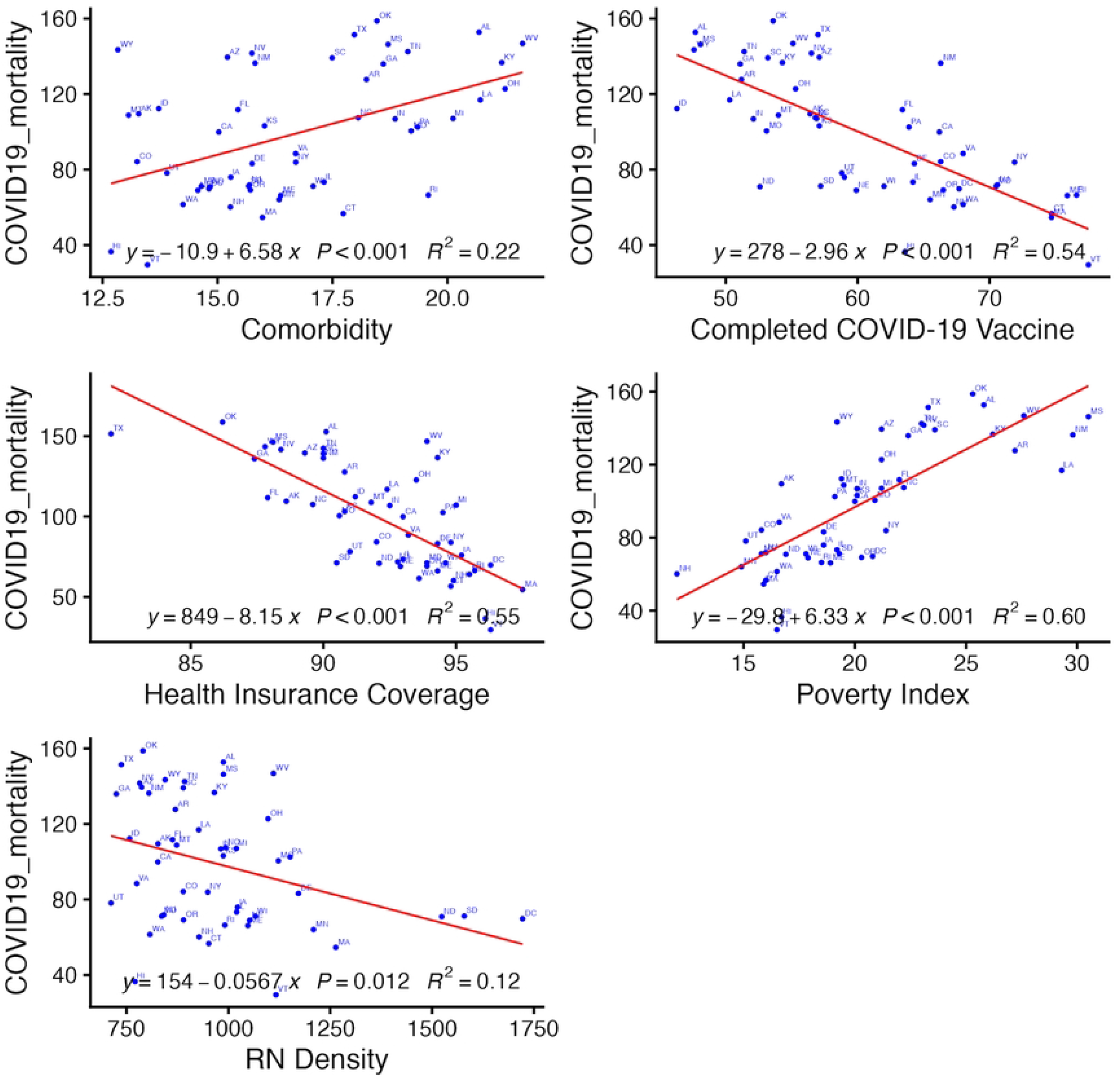
Simple linear regression plots. RN density and all four covariates displayed a statistically significant association with COVID-19 mortality. These plots provide the regression equation for each individual regression, with its p-value and unadjusted R^2^.

### 3.3. Multivariate Linear Regression Results

Subsequently, after controlling for these covariates, the effect of RN density remained statistically significant (B = -0.024, *β*= -0.146, 95% CI: -0.044 to -0.003, p = .024). Overall, the adjusted model explained 86% of the variance of age-adjusted COVID-19 mortality (R^2^ = 0.86, F_(df)_ (5, 45) = 55.67, *p* < .001). The unique contribution of RN density to the explained variance in age-adjusted COVID-19 mortality was around 11% (partial R^2^= 0.1086) in the multivariate model. Health insurance coverage and vaccination against COVID-19 were negatively associated with COVID-19 mortality, while the association was positive for state-level percentage of comorbidities and poverty index (Table 2). All assumptions for the model were satisfied, except DC and four states (namely SD, LA, HI, and WY) were identified as both outliers and influential points with high Cook’s distance (Supplemental information: Fig S1).

**Table 2.**
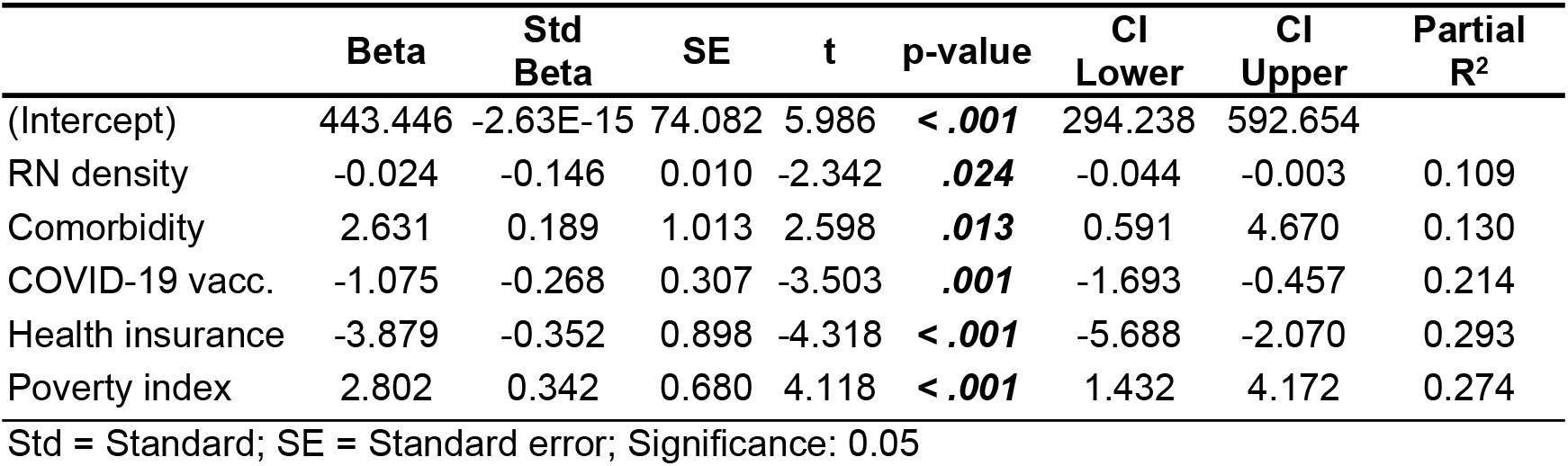
Multiple linear regression results.

Stepwise analysis suggested retaining all the covariates in the final model.

### 3.4. Sensitivity Analysis

Sensitivity analysis was conducted to better capture the influence of these influential states on the multivariate regression model. First, the model was refitted with a log transformation on RN density, which didn’t change the direction of the association between RN density and COVID-19 mortality(B = -26.51, 95% CI: -49.32 to -3.70, p = .024). The residual diagnostics did not indicate any improvement of the influence of these observations, despite the transformation (Supplemental information: Fig S2). Therefore, the linear scale was retained for interpretability.

Furthermore, a model without DC alone, and a third one without DC and all the influential states (SD, LA, HI, and WY) were estimated. Compared with the initial adjusted model, the reduced models showed a better fit, other than the loss of respectively 1 and 5 degrees of freedom (Table 3).

**Table 3.** Sensitivity analysis (Models comparison)

| Model | R<sup>2</sup> | Adj. R<sup>2</sup> | F statistic | df | P value | AIC | BIC | RMSE |
| --- | --- | --- | --- | --- | --- | --- | --- | --- |
| Without influentials | 0.892 | 0.876 | 57.625 | (6, 35) | < .001 | 322.364 | 334.359 | 10.398 |
| Without DC | 0.866 | 0.851 | 56.813 | (6, 44) | < .001 | 406.660 | 420.044 | 12.276 |
| Initial adjusted | 0.861 | 0.845 | 55.674 | (6, 45) | < .001 | 416.155 | 429.678 | 12.475 |
| Log-trans. RN density | 0.861 | 0.845 | 55.670 | (6, 45) | < .001 | 416.158 | 429.681 | 12.476 |
| Population weighted | 0.867 | 0.853 | 58.823 | (6, 45) | < .001 | 425.726 | 439.248 | 12.960 |

df = degree of freedom
Log-trans = Logarithmic transformation

In addition, a slight increase in the magnitude of the effect size for RN density was also observed in both models, respectively (B=-0.033, *β*= -0.17615, 95% CI: -0.056 to -0.009, p-value = .007) and (B= -0.004, *β*= -0.175, 95% CI: -0.067 to -0.014, p-value = .004). The final model was fitted to estimate regression coefficients using weighted least squares, with weight proportional to individual state’s population size, to account for variation across US’ states’ population. The result showed no noticeable difference compared to the previous models (Table 3).

## 4. DISCUSSION

COVID-19 disrupted historical mortality trends in the US, causing the largest increase in annual reported deaths over the last century. The impacts of the pandemic resonated beyond the striking death toll, disrupting the country’s economy and challenging our ability to respond to public health crises. Nurses are the gatekeepers of the nation’s health. Historically, whether in the halls of US Congress or in the hearts of nurse advocates, the urgent need to address the nursing shortage has been a critical debate. Although many investigations have examined how variations in nursing workforce have impacted COVID-19 mortality rates, this relationship remains unsubstantiated at the state level. This ecological study indicates an estimated 24 to 44 decrease in age-adjusted COVID-19 deaths for every 1000 increase in RNs per 100,000 people at the state level, holding constant state-level percentage of chronic health conditions, percentage of the population who completed at least one series of COVID-19 vaccines, percentage of the population with health insurance coverage, and percentage of population who are living below 150% of the poverty threshold.

The regression models provide supportive evidence and detailed information on the magnitude of the observed relationship. A small variance in age-adjusted COVID-19 death across the U.S. states was explained by RN density alone (partial R^2^ = 0.109) as compared to the other covariates (Table 2). Nonetheless, this study provides some supporting evidence of the unique contribution of the nursing workforce on preventing the death of the US population at the state level. Similarly, access to health care insurance and completing at least one series of anti-SARS-CoV-2 vaccines played a critical role in reducing COVID-19 death rates at the state level. Conversely, chronic health conditions and living below 150% of the poverty threshold were associated with higher COVID-19 death rates.

This study paid particular attention to the DC and decided to include it in this analysis. While this decision was beneficial for this analysis, as it helped in increasing the sample size and broadening the study’s scope, it also introduced some expected caveats due to DC’s unique characteristics, making it an outlier among the US states. In this analysis, DC and four U.S. states (HI, SD, MO, WY) were affecting the model’s performance. DC and SD had the highest RN density during the study period. As shown in Fig 4, COVID19 and RN density simple regression plot, these outliers were markedly distant from the other states. The states of HI and WY had the lowest rate of chronic health conditions among US states in 2021. These extreme data points were influencing, but their impact didn’t significantly affect the model’s estimates.

Prior ecological studies that investigated COVID-19 mortality across the US focused on factors such as sociodemographic, comorbidities and regional disparity(34,35). An observational analysis that analyzed factors associated with cross-state variations in infection and mortality rates, found lower poverty rate, higher education, access to quality healthcare, and higher vaccination rates were associated with lower COVID-19 infection and death rates(36). While states with larger percentages of the population identifying as Black or Hispanic were associated with higher cumulative death rates(34,36). One study found no statistical association between higher public health personnel per capita with a decrease in COVID-19 infections or death rate, at the state level(36).

Most studies that explicitly assess the relations between COVID-19 and healthcare workers (HCW) focus on the direct impact of the pandemic on that population. Globally, between 2020 and 2021, WHO reported an estimated 6,643 COVID-19 deaths among HCW(37). In the US, by April 2021, an estimated 3,607 deaths within HCW was reported, with the highest death toll among nurses(32%)(38). Other impacts of the pandemic on HCW were reported, including stress, anxiety, depression, and burnout (39). An estimated 5% of the nursing workforce left the profession because of the stress load they experienced during the COVID-19 pandemic(40).

One cross-country analysis that investigated the association between healthcare workforce and health outcomes found a higher density of nursing personnel was associated with a lower level of COVID-19-associated excess deaths per 100,000 people(41).

This study uses reported data from reputable national databases. Despite a small sample size(n=51), this analysis used linear regressions to predict state-level age-adjusted COVID-19 mortality with RN density. Our study carries multiple limitations including a small sample size and the cross-sectional approach that restricts its generalization to a broader extent.

Furthermore, while these data were from reputable sources, these cumulative estimates may carry some misestimation. Most importantly, the inherent, ecological nature of this study is one of the key limitations, restricting the findings from being interpreted from a population-based context as opposed to a more disaggregated, local, or individual level.

Despite the above limitations, this study’s findings are not trivial. Actionable policies-driven interventions can be implemented by building on this study’s findings. First, our study could be of benefit to public health policymakers for planning and developing healthcare systems with a better ability to respond to emergencies such as COVID-19 pandemic, at the state level. At the federal level, agencies responsible for health resources and services administration should consider incorporating state-level RN density as a key metric for healthcare surveillance and emergency preparedness, to allow adequate RN distribution and prevent avoidable mortality, at the state level. This latter requires additional data to account for disparity in RNs-to-population ratio between administrative subdivisions within an individual state, allowing fair allocation of available resources. Furthermore, the findings underscore the importance of prioritizing investments in the nursing workforce. More efforts are needed to optimize recruitment, retention, and nurses’ overall resilience, especially in geographic areas that are disproportionately more affected by the nursing shortage. Overall, this study highlights the vital role of the nursing workforce in responding to public health emergencies, ensuring the wellness and security of the American people.

## 5. Conclusion

Numerous studies have shown how many Americans with COVID-19 who were admitted to hospitals with an adequate nursing workforce were more likely to survive. This study extends these investigations to the state level where both RN density and reported COVID-19 mortality rates varied substantially. Nurses face numerous challenges in their preparedness and their ability to respond to health emergencies, including staff shortages, insufficient or inappropriate training and resources, and psychological distress. Addressing these challenges requires comprehensive strategies to empower both nurses and state officials. Most importantly, urgent and concrete efforts are needed to overcome the nursing shortages in the US, ensuring equitable resource allocation at all levels. This latter requires joint commitment from governmental, private, and non-profit entities to promote optimal distribution of nurses across the country, with fair and effective incentive systems.

Future studies should extend the scope beyond RNs to include both practical and advanced practice nurses. In addition, this analysis can be further explored at lower administrative divisions, to disaggregate these estimates and unmask existing disparities within individual states. This will also allow us to assess whether these findings persist at lower administrative settings such as counties and municipalities or are simply limited to the ecological context.

Moreover, while we continue to advocate for policy changes at the congressional level that can substantially abate the nursing shortage, a paradigm shift in nursing research, education, and public health is needed to strengthen the nursing workforce’s resilience and readiness to respond to future public health crises.

## Data Availability

All data produced are available online at

https://github.com/lrigan2025/Data_and_Script.git

## Acknowledgements

This research was made possible through the generous support of the University of Florida College of Nursing and the Emerging Pathogens Institute.

## Ethics declarations

This investigation exclusively used deidentified data from publicly available sources. Therefore, Institutional Review Board approval was not needed.

## Data availability

The final data for this analysis and R-script are available on GitHub and can be accessed using the following link: https://github.com/lrigan2025/Covid-19-Mortality-and-Nursing-Workforce-.git

